# Scaling text de-identification using locally augmented ensembles

**DOI:** 10.1101/2024.06.20.24308896

**Authors:** Karthik Murugadoss, Saivikas Killamsetty, Deeksha Doddahonnaiah, Nakul Iyer, Michael Pencina, Jeffrey Ferranti, John Halamka, Bradley A. Malin, Sankar Ardhanari

## Abstract

The natural language text in electronic health records (EHRs), such as clinical notes, often contains information that is not captured elsewhere (e.g., degree of disease progression and responsiveness to treatment) and, thus, is invaluable for downstream clinical analysis. However, to make such data available for broader research purposes, in the United States, personally identifiable information (PII) is typically removed from the EHR in accordance with the Privacy Rule of the Health Insurance Portability and Accountability Act (HIPAA). Automated de-identification systems that mimic human accuracy in identifier detection can enable access, at scale, to more diverse de-identified data sets thereby fostering robust findings in medical research to advance patient care.

The best performing of such systems employ language models that require time and effort for retraining or fine tuning for newer datasets to achieve consistent results and revalidation on older datasets. Hence, there is a need to adapt text de-identification methods to datasets across health institutions. Given the success of foundational large language models (LLMs), such as ChatGPT, in a wide array of natural language processing (NLP) tasks, they seem a natural fit for identifying PII across varied datasets.

In this paper, we introduce locally augmented ensembles, which adapt an existing PII detection ensemble method trained at one health institution to others by using institution-specific dictionaries to capture location specific PII and recover medically relevant information that was previously misclassified as PII. We augment an ensemble model created at Mayo Clinic and test it on a dataset of 15,716 clinical notes at Duke University Health System. We further compare the task specific fine tuned ensemble against LLM based prompt engineering solutions on the 2014 i2b2 and 2003 CoNLL NER datasets for prediction accuracy, speed and cost.

On the Duke notes, our approach achieves increased recall and precision of 0.996 and 0.982 respectively compared to 0.989 and 0.979 respectively without the augmentation. Our results indicate that LLMs may require significant prompt engineering effort to reach the levels attained by ensemble approaches. Further, given the current state of technology, they are at least 3 times slower and 5 times more expensive to operate than the ensemble approach.

## Introduction

The free text clinical narratives from Electronic Health Records (EHRs) can cover a wide variety of patient histories, testing and imaging assessments, diagnoses, treatment plans, and other important medical information. Under the Health Insurance Portability and Accountability Act of 1996 (HIPAA), EHR data can be shared when it is considered to be de-identified^1^. Access to de-identified datasets across multiple healthcare institutions supports various clinical research applications including, but not limited to, creation of patient cohorts^2^, tracking patient journeys^3^ and the development of predictive algorithms^4,5^ (**Figure 1A**).

**Figure 1.**
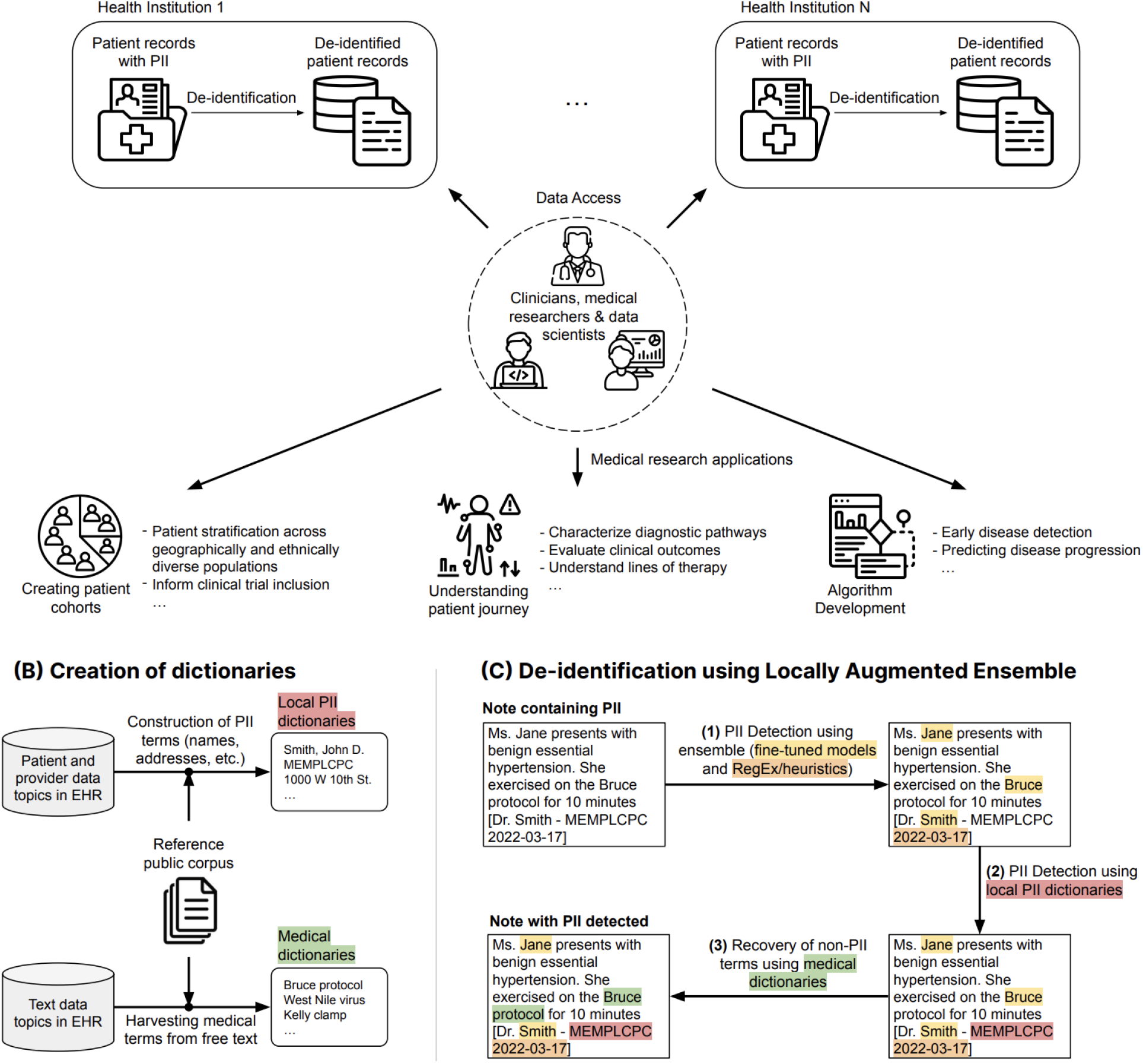
Locally augmented ensemble for de-identification of unstructured text: **(A)** De-identified datasets from multiple health institutions support various clinical research applications. **(B)** Automated framework to create PII dictionaries from health institution specific patient and provider data. **(C)** Runtime application of local dictionaries. (1) The fine-tuned transformer models and regular expressions are run on notes to capture baseline PII (2) any residual PII is detected using the PII dictionaries (e.g. MEMPLCPC - which is an abbreviation of the fictional “Memorial Place Primary Care”). (3) The usage of medical dictionaries helps in untagging “Bruce” in “Bruce protocol” - which was previously incorrectly detected as PII by the fine-tuned models. Overall, the locally augmented ensemble improves the detection of PII and reduces the misclassification of medically relevant terminology. The fine-tuned models and RegEx heuristics module is presented in greater detail in Figure 2 of Best-in-class de-identification paper (Murugadoss et al.)^10^.

The Safe Harbor approach to de-identification under the HIPAA Privacy Rule requires the elimination of 18 categories of identifiers (e.g., patient names, dates of birth, addresses). Various approaches to Safe Harbor de-identification have been developed, including rule-based, traditional machine-learning, deep-learning, and hybrid or ensemble systems^6–9^. Ensemble methods incorporating the transformer architecture, in particular, have demonstrated improved accuracy and robustness in de-identifying data by combining the strengths of multiple approaches^10,11^. These approaches typically leverage regular expressions, heuristics, and the contextual understanding of attention-based deep-learning models to effectively identify PII (personally identifiable information) in unstructured clinical text.

Despite their state-of-the-art performance on public benchmarks and proprietary clinical datasets^10^, ensemble de-identification methods offer ample opportunity for improvement. Even solutions using best performing deep learning models pre-trained on large public corpora may fail to detect out of distribution terms like uncommon names^12^. These pre-trained models may also struggle to identify health institution-specific PII terminology. For instance, satellite clinics affiliated with a major hospital are usually referred to using custom abbreviations which are not released publicly. Another significant challenge arises when de-identification systems, designed to maximize PII detection, inadvertently classify useful information as PII. This issue is non-negligible in medical literature due to the presence of clinical eponyms (e.g., New York Heart Association Class, Bruce protocol, Tuohy needle, and Kawasaki disease)^13^. Such misclassifications compromise the accuracy and reliability of medical data, ultimately reducing its value for research and clinical purposes.

Large language models (LLMs) provide a promising opportunity for tackling the intricacies of clinical text de-identification^14^. Instruction-tuned LLMs are particularly versatile in complex applications, and have demonstrated surprising capabilities across a wide variety of domains and tasks^15^. These models, especially in informal settings, have demonstrated a proficiency in identifying and classifying various entities (such as names, locations and dates) embedded in free text. However, when subjected to established Named Entity Recognition (NER) or information extraction (IE) benchmarks, the current state-of-the-art LLMs fail to outperform supervised fine-tuned models^16–18^. The utilization of LLMs in specialized documents for clinical NER and financial NER have also revealed limitations in its ability to handle domain-specific knowledge and terminology^17,19^. Moreover, throughput is a crucial factor when deploying LLMs, as models with billions of parameters demand increased computational resources and processing time to generate responses for a given input. This poses challenges for their practical application in real-time or resource-constrained environments. LLMs, however, are evolving at a rapid pace with new models, open as well as closed source, being released every month with improved performance. We, therefore, evaluate LLMs on the de-identification task to characterize their performance and understand their capabilities.

In this work, we present the “locally augmented ensemble” approach to address the challenges of text de-identification in a two-step process. First, we introduce an automated framework to create health institution-specific dictionaries of PII as well as medical terminology (**Figure 1B**). Secondly, we discuss the integration of these dictionaries into existing ensemble de-identification systems to detect rare PII elements and institution-specific identifiers and to recover medical vocabulary that has been mis-classified as PII (**Figure 1C**). We demonstrate that our proposed approach enhances the accuracy of clinical text de-identification in real-world clinical datasets from Duke University Health System and Mayo Clinic using a base ensemble fine-tuned using notes from Mayo Clinic^10^. In particular, the ensemble comprises transformer neural networks which are supported by regular expressions and heuristics. Finally, we evaluate LLMs on the de-identification task using the i2b2 2014 De-identification Challenge and the CoNLL 2003 NER dataset and compare these LLMs’ capabilities with our approach^20,21^. We also provide an analysis of inference throughput times and cost estimates.

## Results

### Datasets, evaluation metrics and models

We evaluated the performance of the locally augmented ensemble on data from two geographically distinct healthcare systems. The first is from Duke University Health System and is composed of 15,716 clinical notes. These notes are sampled from a corpus of over 100 million clinical notes corresponding to 250,000 unique patients. The second is composed of 10,000 clinical notes from Mayo Clinic and sampled from over 250 million clinical notes corresponding to 700,000 unique patients. These datasets are referred to as the Duke and Mayo test sets, respectively. They were created using stratified sampling, such that notes were partitioned into groups by type and creation date and sufficient representation from each group was ensured (see **Methods**). As a result, the datasets consisted of various different types of notes such as progress notes, H&P notes, pathology reports, telephone encounters, treatment summaries, etc. spanning from 1990 to 2024. The datasets were manually annotated to create ground truth labels for PII. Each sentence was initially annotated by two different people and conflicting annotations were adjudicated by a third person. Consequently, these datasets are regarded as the gold standard for our evaluation.

De-identification performance was measured using precision, recall and F1-score. Here, precision refers to the proportion of correctly identified PII entities out of all the entities the model identified as PII. Recall refers to the proportion of actual PII entities in the data that the model successfully identified and from a regulatory perspective, high recall must be achieved. The F1-score is the harmonic mean of the precision and recall. Also, we include entities beyond the standard PII identifiers defined by HIPAA (e.g., doctor’s names and clinic locations) as part of our evaluation (see the List of PII entities section in the **Supplementary Information**) given that many healthcare organizations aim to protect the privacy of their care providers as well. Additionally, the reported precision and recall are calculated by distinguishing only between PII and non-PII entities, thereby not penalizing misclassifications within PII categories, such as mistaking organizations for locations (see the Evaluation metrics section in the **Supplementary Information**). In addition to these core performance metrics, we also evaluated the latency and cost of execution.

Finally, we performed comparisons against the best performing^22^ publicly-available open-source Language Learning Models (LLMs) and ChatGPT using two publicly-available datasets. For our analysis, we select the instruction-tuned variants of Llama-3 (Llama-3-8B, Llama-3-70B)^23^ and Mixtral 8x7B^24^, which match GPT-3.5 across various Natural Language Understanding (NLU) and reasoning benchmarks. In addition to open-source LLMs, we also evaluate GPT-3.5 and GPT-4 given their state-of-the-art capabilities and widespread adoption^25^.

We evaluated the LLMs on the i2b2 2014 and the CoNLL 2003 datasets. The i2b2 2014 dataset is a publicly available dataset of clinical documents with annotated PII^20^. This dataset consists of a training set of 792 clinical notes and a test set of 515 clinical notes. The CoNLL 2003 dataset is a benchmark collection for Named Entity Recognition (NER), featuring annotated texts with entities like names and locations, and comprises a training set of 946 news articles and a test set of 217 news articles^21,26^.

The source code to run the large language models on the CONLL 2003 and the I2B2 dataset is available at https://github.com/lumenbiomics/udeid_llm.

### Locally augmented ensembles achieve best-in-class performance on real-world clinical datasets

We evaluated the end-to-end de-identification system on both the Duke and Mayo test sets (see **Table 1**). For the Duke test set, the precision was 0.982 (95% CI: 0.981, 0.982), recall was 0.996 (95% CI: 0.996, 0.996), and the F1-score was 0.989 (95% CI: 0.989, 0.989). For the Mayo test set, our approach achieved a precision of 0.979 (95% CI: 0.978, 0.980), a recall of 0.995 (95% CI: 0.995, 0.995), and an F1-score of 0.987 (95% CI: 0.987, 0.987). As can be seen, the overall levels of precision and recall between the two healthcare systems are similar. A deeper analysis of the types of false negatives is presented in **Supplementary Table 1**. Additionally, we assessed the performance of our system in the absence of specific components namely, patient and provider dictionaries that are derived from the EHR and clinical terminologies. We discuss these components in greater detail in the following sections.

**Table 1.** An evaluation of de-identification models on the Duke and Mayo test sets.

| **Method Configuration** | **Duke** |  |  | **Mayo** |  |  |
|---|---|---|---|---|---|---|
|  | **Precision** | **Recall** | **F1-Score** | **Precision** | **Recall** | **F1-Score** |
| Ensemble without local augmentation (Baseline) | 0.971 | 0.989 | 0.980 | 0.972 | 0.995 | 0.983 |
| Baseline with Structured PII dictionaries | 0.971 | 0.996 | 0.983 | 0.972 | 0.996 | 0.984 |
| Baseline with medical dictionaries | 0.982 | 0.989 | 0.985 | 0.979 | 0.995 | 0.983 |
| Locally augmented ensemble | 0.982 | 0.996 | 0.989 | 0.979 | 0.996 | 0.987 |

### Using dictionaries for PII detection

Our ensemble approach uses dictionaries of PII terms from patient and provider topics in the EHR to detect PII in clinical text, including multi-word terms like clinic names and locations, as well as full names of physicians and patients. Additionally, a patient-specific dictionary is created, mapping patient IDs to names and unique identifiers. The incorporation of these dictionaries primarily aims to enhance the performance of the existing transformer models, especially by capturing health institution-specific PII terms and rare patient or physician names, which are less likely to appear in public literature that inform pre-trained language models.

When the structured PII dictionaries are utilized, we observe an increase in recall from 0.989 to 0.996 on the Duke test set and from 0.995 to 0.996 on the Mayo test set (see **Table 1**). Of note, the baseline recall in the Mayo test set is higher than the Duke test set since the models were fine-tuned with samples from Mayo. As expected, the precision remains unaffected because of the quality (i.e no ambiguous terms) of the PII dictionaries. We also provide several examples of the kinds of entities that were missed by the ensemble but were correctly captured using the PII dictionaries (see **Table 2**). In the absence of structured PII support for the evaluation on the Duke test set, it is noteworthy that the transformer models, trained exclusively using Mayo Clinic documents, were able to generalize to the language and structure of clinical notes from Duke and achieve a high recall.

**Table 2.** Examples of (fictional) sentences with PII detected using structured PII dictionaries: Sentences from the Duke and Mayo test sets with PII (underlined) that were not detected by the ensemble but were captured through the PII dictionaries..

| **PII Entity Type** | **Example Sentence with PII** (fictional) **term** |
|---|---|
| Patient Name | Born denies any PND or claudication symptomatology. |
| Provider Name | GH8715 Sylvia Ayonghe Ndu MD 05/13/2018 |
| Patient Address | Transfer: 40 CROSSBEAK RD, ROCHESTER, MN |
| Clinic Location | Answer: 2000 Stardust Dr. / Order Specific Question: |
| Organization | Referral Case 00-0012 MSHJ HEALTH CARE, Minneapolis |
| Organization | 3/17/2019 4:00 PM John B. Smith, MD DURPLCPC DUKE CHILD Patient Instructions Increase Amlodipine to 10 mg once daily |

### Recovery of medically relevant false positives

The ensemble approach occasionally misclassifies medically relevant terms as PII. To address this issue, we established a dictionary of medical terminology harvested from public datasets and clinical text. Using this medical dictionary, we observed that the precision of the unstructured de-identification software increased from 0.971 to 0.982 on the Duke test set and from 0.972 to 0.979 on the Mayo test set while the recall remains the same (see **Table 1**). Although we observed an increase in precision on the test sets, it is important to note that these test sets were created primarily for measuring the de-identification recall. More specifically, the selection of documents for the test sets was optimized for high PII content (see Methods). Therefore, to better quantify the impact of medical term recovery, we measured the number of recovered terms, words detected as PII that also appeared in the medical dictionary, in a random sample of 10M clinical notes from both the Duke and Mayo corpora. We found that 3.5% of terms which were originally labeled as PII in the Duke test set are recovered and while % of terms in the Mayo test set are recovered (see **Table 3**). The percentage of recovered entities in the Mayo test set is lower than that of Duke since the models were fine-tuned on the Mayo test set. We present examples of commonly occurring medical terms across various entity types that were correctly captured using the medical dictionary (see **Table 4**).

**Table 3.** Number of recovered terms: Across the Duke and Mayo 10M notes dataset, we determine the number of recovered words for each entity type amongst the total number of words detected as PII.

| **PII Entity Type** | **Duke** |  | **Mayo** |  |
|---|---|---|---|---|
|  | **Words detected as PII** | **Recovered Words (%)** | **Words detected as PII** | **Recovered Words (%)** |
| Age | 5,572,744 | 9,627 (0.2%) | 2,608,541 | 11,495 (0.44%) |
| Alphanumeric Identifier | 2,505,651 | 37,902 (1.5%) | 66,226,790 | 5,027 (0.007%) |
| Date | 44,117,044 | 693,847 (1.5%) | 47,810,849 | 98,188 (0.21%) |
| Email | 17,819 | 0 (0.0%) | 4 | 0 (0.0%) |
| IP Address | 48 | 0 (0.0%) | 376,083 | 0 (0.0%) |
| Location | 9,244,820 | 1,207,556 (13.1%) | 6,983,875 | 373,765 (5.3%) |
| Name | 31,785,298 | 531,716 (1.7%) | 56,515,604 | 286,800 (0.5%) |
| Organization | 8,260,065 | 1,191,949 (14.4%) | 4,708,642 | 609,347 (12.9%) |
| Pager | 136,491 | 20,966 (15.4%) | 294,881 | 420 (0.1%) |
| Phone number | 1,799,581 | 6,364 (0.3%) | 3,894,765 | 606 (0.015%) |
| Vehicle Plate | 2,899 | 0 (0.0%) | 29,327 | 0 (0.0%) |
| Website | 41,673 | 1 (<0.1%) | 120,816 | 0 (0.0%) |
| Zip Code | 660,947 | 7 (<0.1%) | 564,874 | 1 (<0.1%) |
| **All** | 104,145,080 | 3,699,935 (3.5%) | 190,199,105 | 1,385,649 (0.7%) |

**Table 4.** Examples of highly prevalent false positives that are recovered using the medical dictionaries: Medical terms and example sentences from the Duke and Mayo 1M notes datasets that were originally detected to be PII. “# Instances Recovered” represents the number of times that the medical term was labeled as PII and subsequently recovered using the medical DB.

| **Medical Term (Type)** | **Instances Recovered (across Duke and Mayo sets)** | **PII Entity Type** | **Example Sentence with recovered medical term** (medical term is underlined, words originally detected as PII are in **bold**) |
|---|---|---|---|
| 3/16” needle (surgical instrument) | 8,404 | Date | • BD INSULIN PEN NEEDLE UF MINI 31 gauge x **3/16"** needle USE UTD QID |
| AAP Clinical Practice Guideline (Clinical practice) | 11,736 | Organization | Blood pressure percentiles are 90 % systolic and 53 % diastolic based on the 2017 **AAP** Clinical Practice Guideline. |
| BASAGLAR KWIKPEN (injection) | 2,986 | Name | • [DISCONTINUED] **BASAGLAR KWIKPEN** U-100 INSULIN pen injector (concentration 100 units/mL) |
| Bruce Protocol (clinical test) | 845 | Name | The patient exercised on standard **Bruce** protocol for 5 minutes and 10 seconds and Achieved target heart rate. |
| FK506 (drug) | 16,713 | Alphanumeric Identifier | No results for input(s): **FK506** in the last 168 hours. |
| NAD (clinical state; “No Apparent Distress”) | 4,970 | Location | General: Patient sitting up in bed in **NAD**. |
| St. Jude (surgical device) | 2,632 | Location | Severe MR and mod TR s/p MVR (29mm **St. Jude**) and TV repair 1/8 |
| MyChart (software application) | 45,439 | Organization | She is to contact the clinic through **MyChart** with major concerns. |
| 0630 (time) | 19,281 | Pager | Inpatient Schedule: Breakfast (**0630**) |

### Locally augmented ensembles outperform large language models on public datasets

Our evaluation involved zero-shot and one-shot prompting techniques for the LLMs^27^. For the I2B2 2014 test set, we found that the Llama-3-70B model is the best performing method among the LLMs, with a precision of 0.89, recall of 0.83 and F1-score of 0.85 (see **Table 5**). By contrast, for the CONLL 2003 test set, the best performing LLM approach was the one shot prompting strategy with GPT-4, which has a lower precision of 0.79, a recall of 0.96, and an F1-score of 0.87. We observed a total of 298 false negative errors with one shot prompting with GPT-4. Locations were the most prevalent entity type with 117 errors (50.6%). Examples of locations that were missed are the abbreviation “U.S.” and the continent “Europe”. Following locations, names constituted the second most significant error category comprising 98 false negatives (32.9%). Personal names that occurred as part of tables were frequently missed as well as infrequent names, such as “Suharto” and “Xue”. False negatives in organizations occurred in 83 instances (27.8%) with errors in terms such as “Reuters” and “Newsroom” (see the Examples of ChatGPT-4 errors section in the **Supplementary Information** for examples of notes and their outputs from GPT-4). On both the I2B2 and CONLL datasets, the ensemble achieves the best performance among all LLMs with a precision of 0.96, recall of 0.99 and F1-score of 0.97 on the I2B2 test set, and a precision of 0.95, a recall of 0.97, and an F1-score of 0.96 on the CONLL 2003 test set.

**Table 5:** Performance of de-identification methods on the i2b2 2014 test set and the CONLL 2003 test set. The precision, recall and F1-score for each LLM/method is described. We did not evaluate GPT-3.5 and GPT-4 on the I2B2 dataset due to data use restrictions.

|  | **I2B2** |  |  | **CONLL** |  |  |
|---|---|---|---|---|---|---|
| **Method** | **Precision** | **Recall** | **F1-Score** | **Precision** | **Recall** | **F1-Score** |
| Llama-3-8B | 0.79 | 0.75 | 0.77 | 0.88 | 0.81 | 0.84 |
| Llama-3-70B | 0.89 | 0.83 | 0.85 | 0.87 | 0.85 | 0.86 |
| Mixtral 8x7B | 0.72 | 0.60 | 0.65 | 0.76 | 0.72 | 0.74 |
| GPT-3.5 Zero Shot | N/A | N/A | N/A | 0.86 | 0.81 | 0.83 |
| GPT-3.5 One Shot | N/A | N/A | N/A | 0.88 | 0.76 | 0.81 |
| GPT-4 Zero Shot | N/A | N/A | N/A | 0.81 | 0.95 | 0.87 |
| GPT-4 One Shot | N/A | N/A | N/A | 0.79 | 0.96 | 0.87 |
| **Locally augmented ensemble (ours)** | **0.96** | **0.99** | **0.97** | **0.95** | **0.97** | **0.96** |

Next, we evaluated the runtime processing throughput of different LLM methods in comparison with our system. We performed this evaluation on the 217 documents present in the CONLL 2003 test set. We observed that our approach requires 0.10 seconds per document while the Llama-3-8B is the second fastest approach with 0.29 seconds per document (**Table 6**). We also provided cost estimates per document for each method. We calculated the runtime cost of open source LLMs using the per hour rates of the 8 x L4 GPU VM (Graphical Processing Unit Virtual Machine) on the Google Cloud Platform, while the exact GPT costs were determined using OpenAI’s pricing rates (See the Cost estimation section in the **Supplementary Information** for more details). For zero shot and one shot prompting in GPT-3.5 and GPT-4, the time elapsed is inclusive of the network latency in the round trip API call. We observed that the per-document time for one shot is lower than zero shot despite the one shot prompt being longer than the zero-shot prompt. However, the longer one shot prompt results in increased costs as compared to zero shot prompting - $299 vs $196 per 10,000 documents for GPT-4.

**Table 6:** Time elapsed for different methods on the CONLL 2003 test set. The total time for returning the named entities for 217 documents in the test set is provided along with the time in seconds per document. * Costs for GPT-3.5 and GPT-4 are computing using OpenAI’s pricing rates. ^#^ The fastest running LLM is the Llama-3-8B model, which is 3 times slower than the locally augmented ensemble ^+^ The cheapest LLM is the GPT-3.5, which is 5 times more expensive than our approach

| **Method** | **Hardware Requirements** | **Total time elapsed for 217 documents (s)** | **GPU-Time (s)** | **Time per document (s)** | **Estimate of compute cost per 10K documents** |
|---|---|---|---|---|---|
| Llama-3-8B | 8 x L4 GPU VMs | 63.21 | 505.68 | 0.29<sup>#</sup> | \$7 |
| Llama-3-70B | 8 x L4 GPU VMs | 311.72 | 2493.76 | 1.43 | \$32 |
| Mixtral 8 x 7B | 8 x L4 GPU VMs | 106.22 | 849.76 | 0.49 | \$10 |
| GPT-3.5 Zero Shot | 1 CPU VM | 1323.78 | N/A | 4.75 | \$5<sup>*+</sup> |
| GPT-3.5 One Shot | 1 CPU VM | 668.36 | N/A | 3.08 | \$5<sup>*+</sup> |
| GPT-4 Zero Shot | 1 CPU VM | 2778.06 | N/A | 12.80 | \$196* |
| GPT-4 One Shot | 1 CPU VM | 1725.97 | N/A | 7.95 | \$299* |
| **Locally augmented ensemble (ours)** | **1 x L4 GPU VM** | **21.70** | **21.70** | **0.10** | **< \$1** |

## Methods

In this section, we outline the methodology employed for creating the gold standard test sets which are used to assess the de-identification performance improvements due to PII dictionaries and recovery of medical terms. We then detail the development and implementation of PII dictionaries and also describe the process for compiling a comprehensive dictionary of medical terms. Lastly, we discuss the testing of unstructured text de-identification using large language models (LLMs).

### Sampling process and creation of gold standard test sets

The Duke and Mayo test sets comprise diverse document types that represent the heterogeneity of clinical notes present in the respective EHR systems. We performed stratified sampling across different types as well as across time to ensure diversity. Specifically, we created multiple document groups with each group representing a unique pair of temporal range and document type. For example, one such group is *Progress Notes* in the time range *2018 to 2021*.

We considered a minimum sample size of the test set to be 10,000 documents (satisfying a 95% confidence level and 1% margin of error in representing all clinical notes). For each <temporal range,type> group, we computed the average number of words. We then determined the adjusted number of documents for each group by multiplying the number of documents with the average number of words. This increases the sampling weight for groups that contain longer documents. Furthermore, highly prevalent document types, such as appointments and medication comments, that contain short strings are suppressed. As an example, in the Duke corpus, there are 12.7M questionnaire responses (answers provided by patients to sets of predefined questions from the health system) from 2021-2022 with 1.99 words per response. In comparison, there are only 3.7M progress notes written in the same time period. However, the progress notes are much longer, with 794 words per note. As a result, progress notes were assigned a higher adjusted count than questionnaire responses (10.8M vs 4.0M) and sampled at a higher rate in the test set.

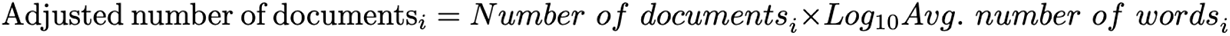

where *i* refers to a year-type group.

We then determined the number of documents to sample for each group:

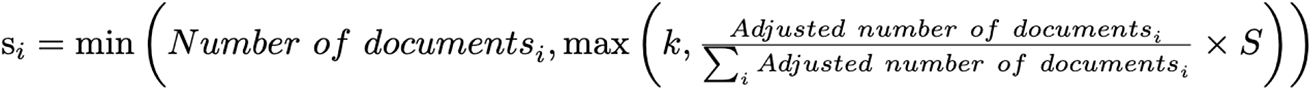

where *s_i_* is the number of documents to sample in the *i*th group, *S* is the minimum sample size (in our case, S=10,000), and *k* is the minimum number of documents to sample for each group. We set *k* to 10, such that every group is comprised of no less than 10 documents unless the bucket itself has fewer than 10 documents. The latter situation can occur for specific (rare) note type-year combinations (e.g. in the Duke corpus, there are only 3 nursing notes present written between 1994-1997, and all 3 notes are included in the test set).

Given the required number of documents to sample per group, the sampling process was guided by the two additional criteria. First, documents from distinct patients were prioritized documents from the same patient. Secondly, documents that contain PII are preferred. A document is deemed to contain PII if it contains either the patient name, address, phone number or email address of the associated patient (taken from the demographics table). Based on this criteria, we observed that approximately 96% of all sampled notes contained instances of PII.

### Using dictionaries for PII detection

To support the ensemble of models, we used dictionaries of PII terms created from the available patient and provider EHR topics as an additional PII detector. Specifically, we created dictionaries of clinic names, clinic locations, physician names, patient names and patient addresses. To support the removal of generic or ambiguous terminology from our dictionaries, we curated a large corpus (referred to as “scientific corpus”) comprising 46 million documents from Pubmed, Clinical Case Reports and the Directory of Open Access Journals (DOAJ). Phrases with an occurrence count greater than 1 million in the scientific corpus (e.g., “Primary Care” or “Radiology Department”) were removed from the PII dictionaries.

In addition to the aforementioned PII dictionaries, we also created a patient specific dictionary of names and unique identifiers. From the patient EHR topic, we created a mapping from each unique patient ID to the first, middle and last name as well as any unique identifiers of the corresponding patient (e.g., patient ID or medical record number). Patient names that correspond to common words (e.g., “Max”, “Will”, or “Green”) were removed if their occurrence count in the scientific corpus exceeded 1 million. We note that this exclusion rule has the side effect of dropping very common names such as “John” or “Mike”. However, these common names are consistently detected by the neural network components of our ensemble and their exclusion from the patient specific dictionaries does not impact overall de-identification performance. On the other hand, a rare name such as “Ndu” may not be detected by the models and its inclusion in the dictionaries improved the overall detection capability.

### Recovery of medically relevant terms

Medically relevant terms, in particular clinical eponyms, may be incorrectly classified as PII (e.g., the word “Bruce” in *Mr. Smith exercised for 4 minutes on the <u>Bruce</u> protocol*). The subsequent transformation of these terms can impact downstream interpretability and analysis (e.g., When replaced by random surrogate values, the sentence becomes *Mr. Johnson exercised for 4 minutes on the <u>Conley</u> protocol* which is incorrect and misleading). We addressed this problem by creating a dictionary of medical terminology from the Duke University Health System and Mayo Clinic corpora. If a term detected as PII is present in this dictionary, then it is deemed medically relevant and is retained as is without any transformation (i.e. it is treated as non-PII).

The dictionary leverages publicly available datasets from the SNOMED^1,28^, Food and Drug Administration (FDA)^29^, and National Cancer Institute (NCI)^30^ databases. Additionally, we developed an approach to harvest medically relevant entities from unstructured clinical text, thereby augmenting the public databases (**Figure 2**). Specifically, we selected multi-gram phrases from clinical text that are also present in public scientific literature (e.g. “Bruce protocol” vs “Bruce jumped”) and are present in a minimum of 10 patients. Additionally, we checked against the structured PII dictionaries (i.e. patient demographics and provider information) to ensure that the entire multigram phrase itself is not PII (e.g. physicians who are prolific publishers). Taken together, a total 460,000 medically/biologically meaningful terms comprised the final medical dictionary used as part of the de-identification system.

**Figure 2.**
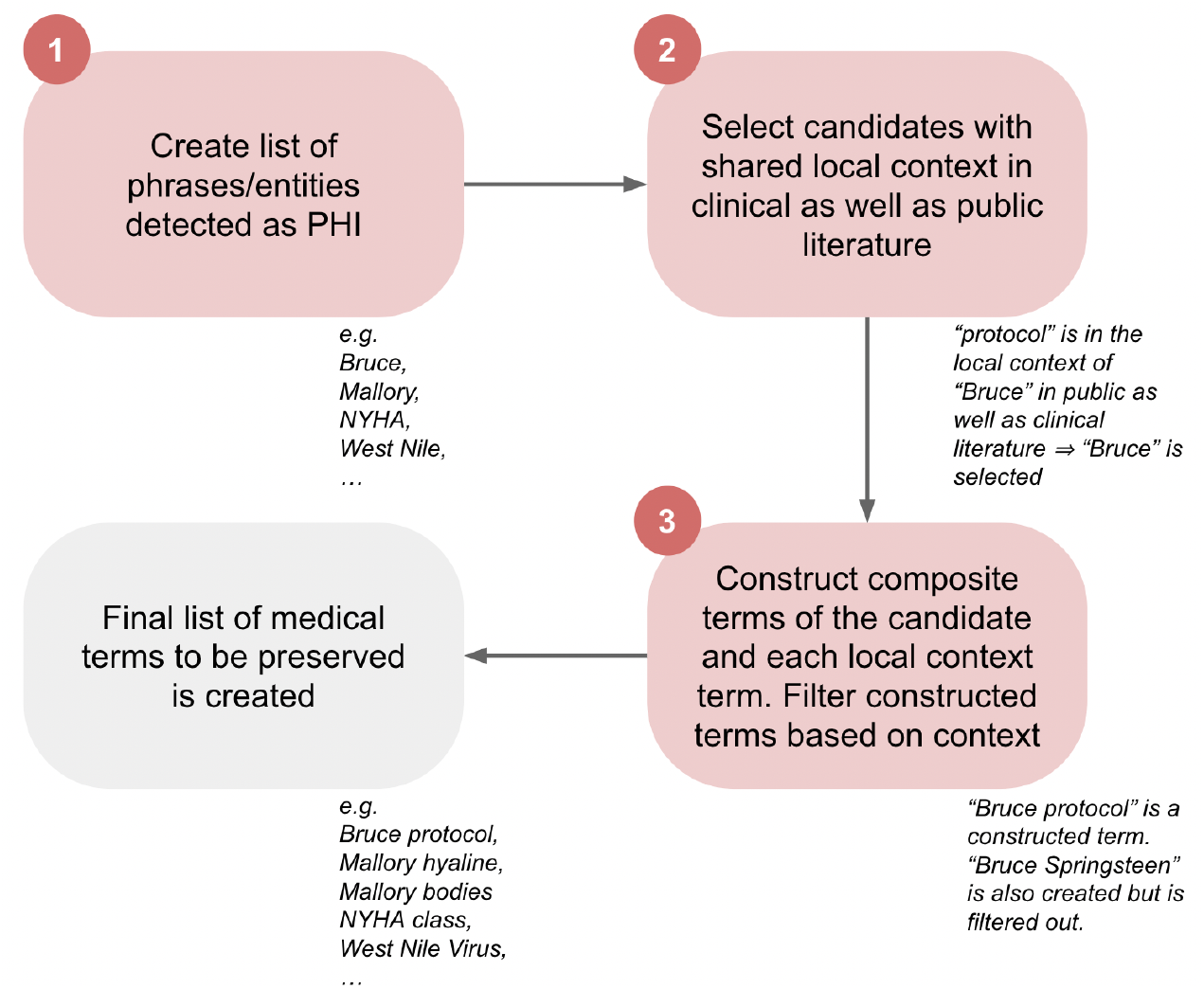
Expanding the medical dictionary: An overview of the process for expanding the dictionary of medically relevant terms that are retained in the unstructured data.

In addition to creating a medical dictionary, we also employed regular expressions to identify potential false positive terms that are mentioned in a consistent and well-defined syntax. For instance, genetic variations are typically represented with a “g.“ followed by a number and the actual nucleotide change (e.g. g.7578395G>C). Therefore, we can employ a specific regular expression (“g\.[0-9_+-]{5,}[TGCA]+(>)?[TGCA]+”) to detect such instances as part of false positive recovery. Words that are detected as PII by unstructured de-identification will be compared with these regular expressions. If such words match the regular expressions in a case insensitive manner, they will be deemed as non-PII. The overall process for recovery of medically relevant terms is shown in **Figure 3**.

**Figure 3.**
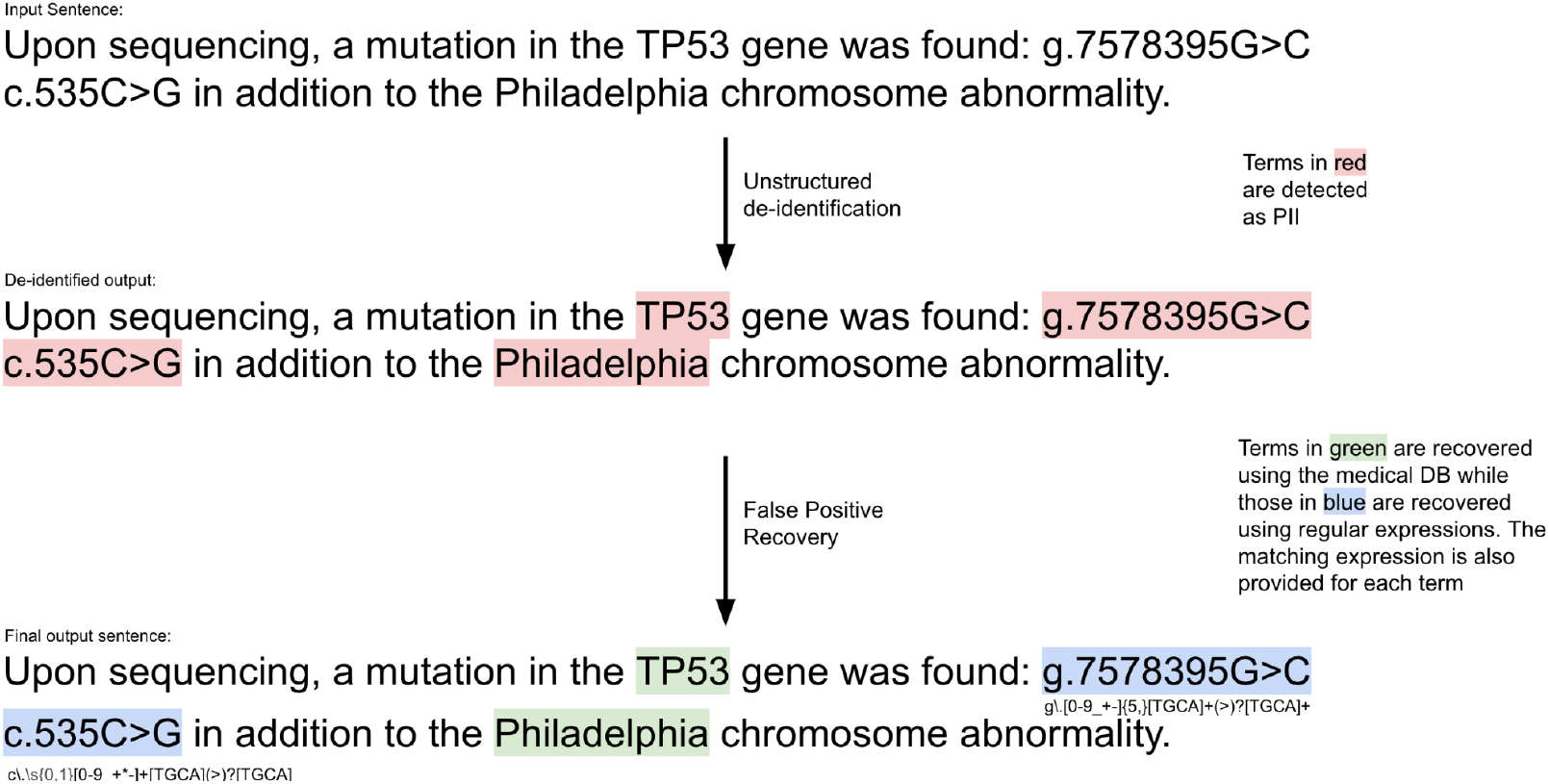
Recovery of false positives during unstructured de-identification: A combination of dictionaries and regular expressions are employed to identify words incorrectly detected as PII. “Recovery” here refers to the process of labeling words that were originally PII as non-PII.

### Unstructured De-identification using Large Language Models

LLMs were evaluated using zero-shot and one-shot prompting. In both scenarios, we instructed the LLMs to return output as a JSON dictionary with keys as entity types and a list of values for each corresponding PII term (**Figure 4**). The raw clinical text from each input document was included as part of the LLM prompt without any modifications. We applied different instruction prompts for different LLMs and included special tokens in the prompt string in accordance with the instructing tuning dataset of each LLM (**Supplementary Table 2**). For GPT-3.5 and GPT-4, each request to the LLM was performed as a separate session to ensure that PII prediction occurs independently on each note. The temperature is an important hyperparameter in natural language generation tasks that controls the creativity of language models and the randomness of the generated text^31^. We set the temperature to zero so that queries were reproducible. After obtaining the response of the language model, a case-insensitive search for performed to identify PII terms in the original text (e.g., if both “John” and “john” occur in the text, only one of them is likely to be returned in the JSON response of the LLM).

**Figure 4.**
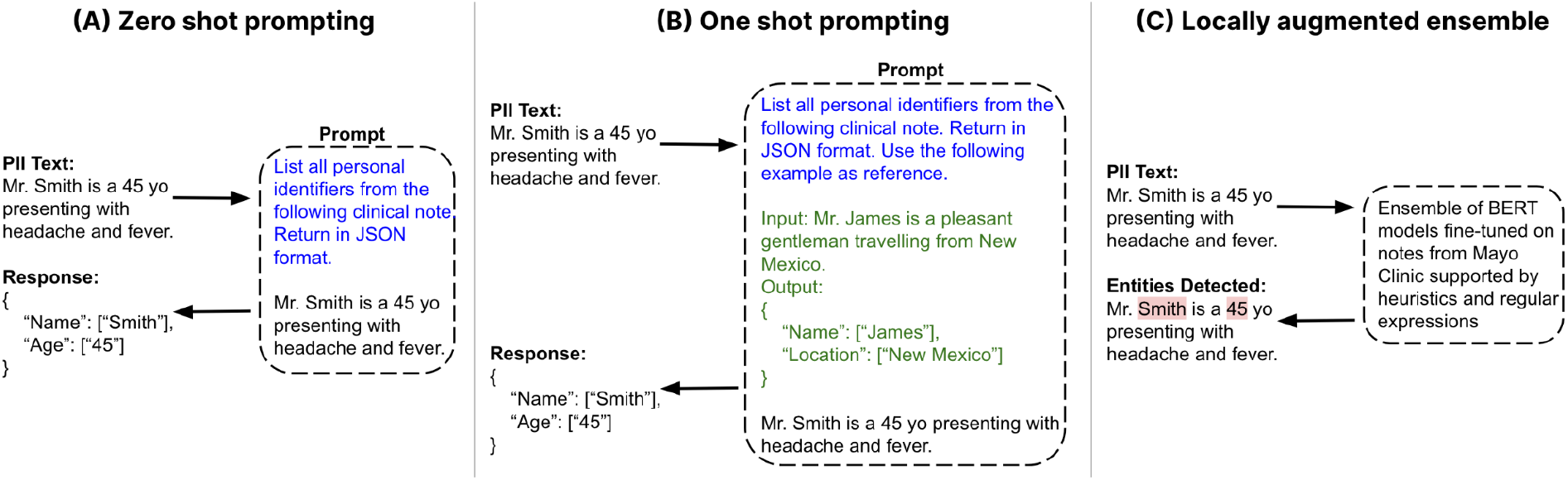
A comparison of de-identification using LLMs and our framework: Depiction of (A) zero-shot and (B) one-shot prompting for de-identification using LLMs. (C) Our methodology. The prompt provided for zero shot and one shot prompting is an example. The actual prompt used in our experiments is provided in Supplementary Table 3.

## Discussion

There is an emerging trend in clinical studies to substitute fine-tuned models with LLMs^32–37^. LLMs can improve patient care by assisting in patient communication through translations and summaries, and simplifying documentation tasks by converting between unstructured and structured information. In medical research, LLMs can distill and summarize research findings, potentially accelerating the integration of new knowledge into clinical practice. Furthermore, in medical education, LLMs are expected to shift the focus towards critical thinking and reasoning, complementing traditional methods of learning. These diverse workflows are supported by the development of numerous open-source LLMs and low-latency API providers. Despite these promising applications and rapid development, the ability of LLMs to detect PII (in zero-shot or one-shot setting) with low false negative rates remains to be demonstrated. Furthermore, even if a highly performant LLM were developed, its compute and cost requirements would be challenging to overcome, especially since processes like de-identification require secure and controlled compute environments (preventing the use of public APIs). One avenue worth considering is the prospect of fine-tuning LLMs, which can help refine their capabilities for specific downstream tasks such as information extraction or de-identification. However, fine-tuning a billion-plus parameter model requires substantial compute requirements. Although compute-friendly implementations such as Parameter Efficient Fine-Tuning (PEFT) may offer a path forward, the benefit of fine-tuning an LLM versus a simpler model (such as BERT or RoBERTa) is still unclear^38,39^.

Beyond new model architectures, another opportunity to improve de-identification performance is by leveraging the syntactic structure and formatting present in clinical documentation. In particular, unstructured text in EHR is increasingly integrated with semi-structured templates (e.g., tables, forms etc.) for consistency and uniformity in clinical documentation. These templates are inherently complex due to inconsistent formatting, vast variability in template designs, and the constant evolution of templates used, which can lead to complications in data retrieval. Despite these challenges, leveraging templatized content from clinical notes offers significant benefits, such as reduced annotation needs for NER models and greater efficiency in model development and validation. Additionally, isolating templatized content reduces the amount of unstructured text processed by deep neural networks, thereby speeding up the overall runtime of tasks like NER and information extraction.

Overall, our investigation demonstrates that a locally augmented ensemble framework for text de-identification achieves best-in-class performance against large and diverse real-world clinical datasets from Duke University Health System and Mayo Clinic. The integration of local dictionaries enhances both the precision and recall of the de-identification system without having to fine-tune the neural network components of the ensemble. Our work also highlights that transformer models fine-tuned at one health institution (Mayo Clinic) can be deployed at another (Duke University Health System) with minimal effort of local dictionary creation. This paves the way for rapid adaptation of the ensemble to clinical datasets from previously unseen health institutions. Moreover, fine-tuning using institutional data for the specific task of entity recognition (personal identifier entities) only updates the weights of the pre-trained/fine-tuned discriminative models^40^. Transferring these models between health institutions does not carry the risk of outputting memorized data associated with generative/autoregressive models. Lastly, the locally augmented ensemble supports configurability, deployment on commodity hardware and scalability across multiple GPUs. Taken together, our framework enables faster access to diverse federated de-identified datasets hence improving the robustness of medical findings and accelerating medical research and advancement.

## Data Availability

- The 2014 i2b2 dataset and the CONLL 2003 dataset are publicly available datasets subject to signed safe usage for research purposes.
- The Duke and Mayo clinical notes are not publicly available at this time.

## Code Availability

- The source code to run both the open-source large language models as well as the GPT-3.5/GPT-4 API on the CONLL 2003 and the I2B2 dataset is available at https://github.com/lumenbiomics/udeid_llm.
- The source code for the locally augmented ensemble implementation is not available.

## Acknowledgements

We extend our gratitude to the Duke University Health System and Mayo Clinic IRB, whose support facilitated the development of the de-identification methods and their testing on real-world datasets. We also thank the clinical and data teams from Duke and Mayo for preparing and helping us understand the EHR data. Specifically, we thank Dr. Anisha Chandiramani, Dr. Armando Bedoya, Ted Ising, Tracey Hughes, Cathy Vanliew and Josh Weatherman from Duke and Ahmed Hadad, Charles Smith, Roberto Blundo, and Phil Fiero from Mayo. We thank Vineet Agarwal, Krishna Prasad Komarath, Jonathan Bell, Vik Nagjee and the nference de-identification team for their support in method development and data analysis. We thank Purushottam Sinha for discussions and guidance around LLM deployment and execution. Finally, we thank Murali Aravamudan, Ajit Rajasekharan, AJ Venkatakrishnan and Venky Soundararajan for their helpful feedback on the manuscript.

## Author Contributions

Conceptualization, K.M., S.A.; Dictionaries for PII detection, K.M., S.K., D.D.; False positive recovery, K.M., S.K., D.D.; Validation, K.M., S.K.; LLMs – implementation & analysis, K.M., N.I.; Writing – original draft, K.M., S.A.; Writing – review & editing, M.P., J.F., J.H., B.M.;

## Supplementary Information

### List of entities considered as PII

1. Age over 89
2. Phone/fax numbers
3. Email addresses
4. Websites and URLs
5. IP addresses
6. Dates
7. Social Security Number
8. Medical Record Numbers
9. Vehicle/device numbers
10. Account/certificate/license numbers
11. Health plan number
12. Street address
13. City
14. Zip Code
15. Employer name
16. Personal names of patients and family members
17. Provider (doctor/nurse) names *
18. User IDs (of care providers) *
19. Health care organization/facility names *
20. Country *
21. State *

* - Considered as PII for our analysis but not included in HIPAA.

### Evaluation Metrics

To evaluate model performance on the de-identification task, we computed the precision, recall and F1 scores. These were computed as follows:

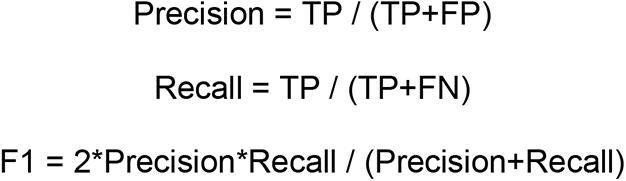

Where,

- TP is the true positive count, the number of words that the ensemble correctly classified as PII,
- FP is the false positive count, the number of words that the ensemble wrongly classified as PII, and
- FN is the false negative count, the number of words that the ensemble failed to classify as PII

Precision refers to the proportion of correctly identified PII entities out of all the entities the model determined to be PII.

Recall refers to the proportion of actual PII entities in the data that the model successfully identified. From a de-identification and patient privacy perspective, maximizing the recall is most important.

The F1 score is the harmonic mean of precision and recall. F1 score is particularly useful when there is an imbalance between precision and recall, providing a single metric that combines both into a balanced evaluation. This is essential for NER systems, which might struggle to balance both precision and recall.

We recognize that alternative performance measures could be utilized, such as specificity, which measures the proportion of true negatives correctly identified. However, specificity is most useful in contexts where both false positives and false negatives have significant consequences and are less relevant for NER systems, such as medical diagnostics. However, in NER, the primary focus is on identifying and classifying entities accurately rather than on recognizing non-entity text as non-entities.

For the precision, recall and F1-Score, 95% confidence intervals were computed using bootstrap resampling with 100 samples.

### Analysis of false negative errors in the Duke Test Set

We analyze the false negative errors based on their risk level for the Duke test set (**Supplementary Table 1**). Here, the first column in this table represents the PII category and the second shows the corresponding risk level for the category while the third column represents the contribution of each category to the false negative error (which corresponds to the residual 0.004 of recall). The error analysis for the Mayo test set is similar to that of Duke and has been presented in our previous publication (See Table 4 of Best-in-Class de-identification paper: https://www.sciencedirect.com/science/article/pii/S2666389921000817).

There are several key findings from this analysis. First, there were six instances of patient names that were not detected by the de-identification system. These were initials, a misspelt name, a nickname and first names that are also common English words (e.g. Will). Taken across 14,631 unique patients, this suggests that the likelihood of leak of patient name is 6/14,631 or 0.0004. There were 10 instances of a leak of the name of a patient’s relative.

Second, there were 56 false negative instances of an alphanumeric identifier. However, upon further analysis, we found that these instances corresponded to slide IDs, accession numbers and physician identifiers that are specific to internal dictionaries and not typically available to the public. None of the instances corresponded to the medical record number, SSN, or account number of a patient.

Third, we found 57 instances of a false negative with dates. Our analysis revealed that none of these dates corresponded to life altering events and were commonly related to date of treatment or procedure.

Fourth, the majority of the false negatives come from clinic location (226 instances) and healthcare organizations (234 instances). These cases typically refer to the site of treatment, physician’s address, etc.

Additionally, we did not come across any false negatives in ages, ZIP codes, websites or email addresses.

**Supplementary Table 1.**
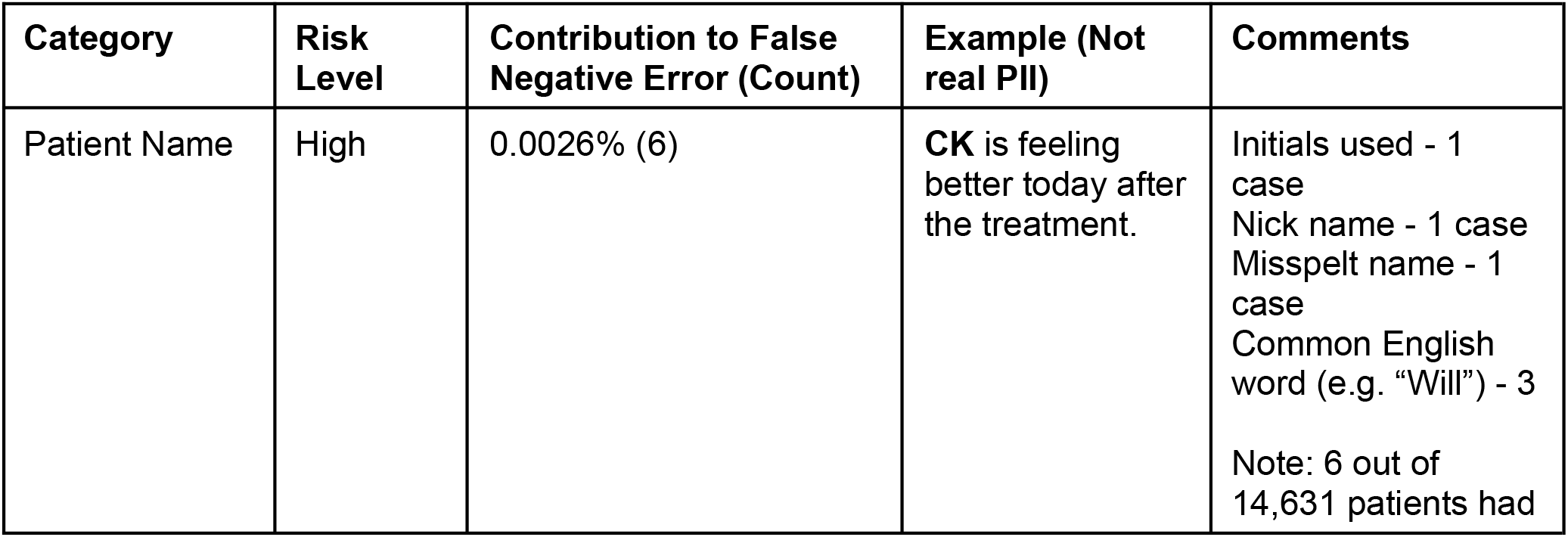
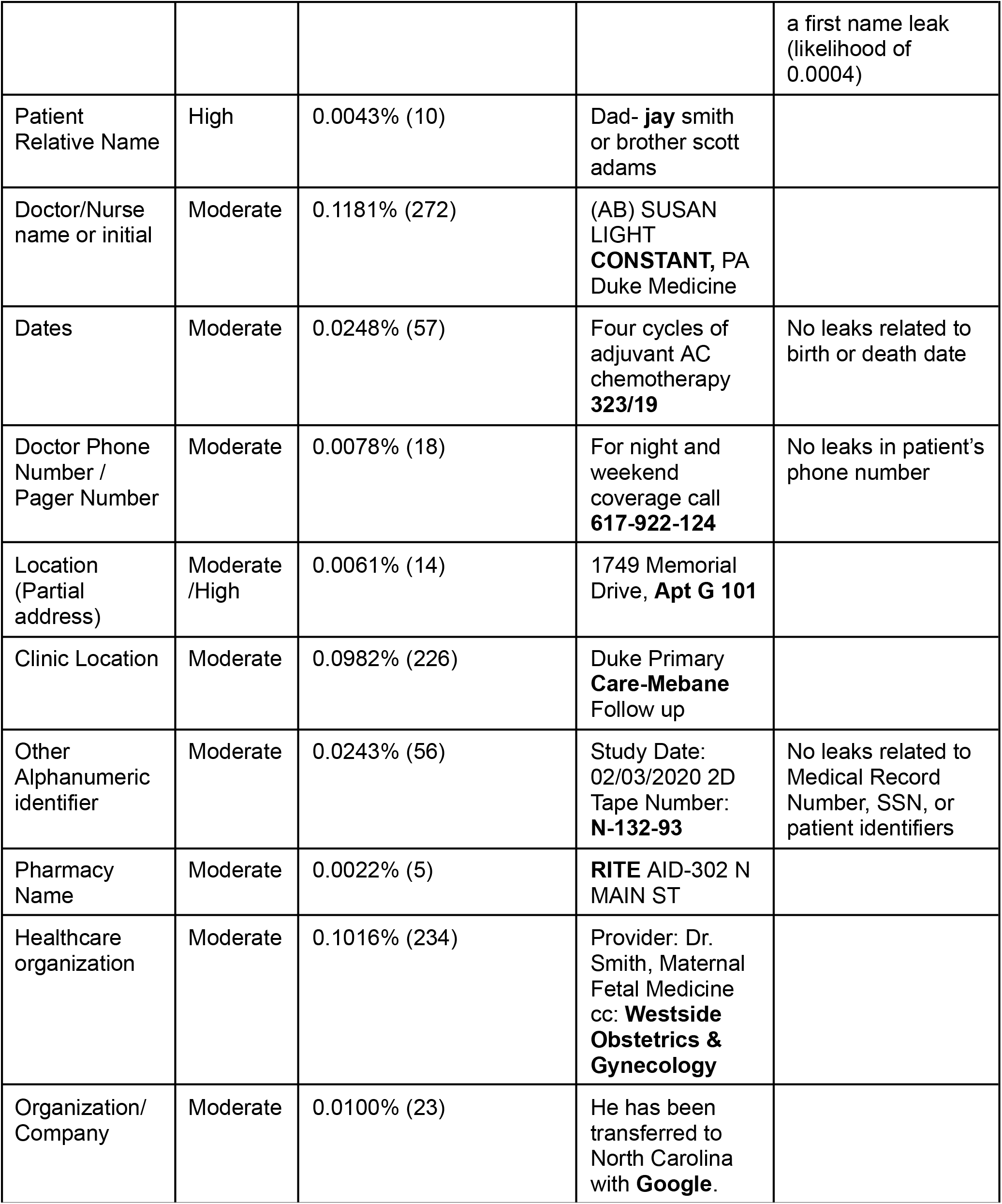
Summary of false negative errors for each entity type: The number of individual false negatives is presented for each entity type along with its percentage contribution to the overall false negative rate of 0.4% (corresponding to a recall of 0.996). Examples are provided for each PII category.

| Category | Risk Level | Contribution to False Negative Error (Count) | Example (Not real PII) | Comments |
|---|---|---|---|---|
| Patient Name | High | 0.0026% (6) | **CK** is feeling better today after the treatment. | Initials used - 1 case<br>Nick name - 1 case<br>Misspelt name - 1 case<br>Common English word (e.g. “Will”) - 3<br>Note: 6 out of 14,631 patients had a first name leak (likelihood of 0.0004) |
| Patient Relative Name | High | 0.0043% (10) | Dad- **jay** smith or brother scott adams |  |
| Doctor/Nurse name or initial | Moderate | 0.1181% (272) | (AB) SUSAN LIGHT **CONSTANT,** PA Duke Medicine |  |
| Dates | Moderate | 0.0248% (57) | Four cycles of adjuvant AC chemotherapy **323/19** | No leaks related to birth or death date |
| Doctor Phone Number / Pager Number | Moderate | 0.0078% (18) | For night and weekend coverage call **617-922-124** | No leaks in patient’s phone number |
| Location (Partial address) | Moderate/High | 0.0061% (14) | 1749 Memorial Drive, **Apt G 101** |  |
| Clinic Location | Moderate | 0.0982% (226) | Duke Primary **Care-Mebane** Follow up |  |
| Other Alphanumeric identifier | Moderate | 0.0243% (56) | Study Date: 02/03/2020 2D Tape Number: **N-132-93** | No leaks related to Medical Record Number, SSN, or patient identifiers |
| Pharmacy Name | Moderate | 0.0022% (5) | **RITE** AID-302 N MAIN ST |  |
| Healthcare organization | Moderate | 0.1016% (234) | Provider: Dr. Smith, Maternal Fetal Medicine cc: **Westside Obstetrics & Gynecology** |  |
| Organization/Company | Moderate | 0.0100% (23) | He has been transferred to North Carolina with **Google**. |  |

For false negatives in dates, we find that 34 of 57 words were missed due to a typo (e.g. “EKG from <u>323/20</u> showed QTc”), 18 words were month and year without the day (e.g. “Chemotherapy was started - <u>11/93</u>”) while the remaining 5 errors were standalone days (e.g. “He was hospitalized till the <u>9th.</u>”).

### Evaluating LLMs

#### Datasets

We downloaded the CONLL 2003 dataset from the Huggingface Datasets portal (https://huggingface.co/datasets/tner/conll2003). The mapping between the entity indices and the entity types were obtained from https://github.com/asahi417/tner. We excluded tags labeled as MISC in our evaluation. The final test set is composed of 217 documents. For our one-shot prompting experiments with GPT-3.5 and GPT-4, we select a single document from the CONLL 2003 training set along with the labels and use this as our one-shot example.

The i2b2 2014 de-identification challenge test corpus is composed of 514 notes. In accordance with our previous work (https://www.sciencedirect.com/science/article/pii/S2666389921000817; see Table 1 and Table 2), we consider HIPAA entities along with provider names and user IDs as relevant entity types for evaluation. In particular, we exclude seasons (e.g., winter and spring) and days of the week from our evaluation.

#### Instruction prompt templates

We use different instruction prompts for different LLMs (**Supplementary Table 2**). Specifically, for the instruction-tuned LLMs (LLama3 and Mixtral), we include special tokens in our prompts so as to mimic the dataset used for fine-tuning. In these templates, only <SYSTEM_PROMPT>, <INSTRUCTION> and <INPUT_TEXT> are placeholders. Also, for GPT-3.5 and GPT-4, we specify the role of the <SYSTEM_PROMPT> as “system”. The remaining instruction in the prompt has the “user” role. Terms such as “<|begin_of_text|>”, “<|start_header_id|>”, “[INST]”, etc. act as markers that indicate where the system prompt and instructions begin or end during the model’s inference process. The same markers were used while instruction-tuning the models and are therefore required at inference time as well.

**Supplementary Table 2.** Instruction prompt templates used for different LLMs.

| **Model** | **Instruction Prompt Template** |
|---|---|
| Llama3 (8B and 70B) | <\|begin_of_text\|><\|start_header_id\|><br>user<\|end_header_id\|><br><system_prompt><br><instruction><br>Input text:<br><input_text><br><\|eot_id\|><\|start_header_id\|>assistant<\|end_header_id\|><br>Output JSON Response: |
| Mixtral 8x7B | <s> [INST]<br><system_prompt><br><instruction><br>Input text:<br><input_text><br>[/INST]<br>Output JSON Response: |
| GPT-3.5/GPT-4 Zero shot | <system_prompt><br><instruction><br>Input text:<br><input_text><br>Output JSON Response: |
| GPT-3.5/GPT-4 One shot | <system_prompt><br><instruction><br>Input text:<br><one_shot_example_input_text><br>Output JSON Response:<br><one_shot_example_response><br>Input text:<br><input_text><br>Output JSON Response: |

#### Prompt engineering

In this section, we present the different prompt strings used for testing the LLMs. The results presented in the paper correspond to the best performing prompt on GPT-4 (Prompt #4 in **Supplementary Table 3**).

Of note, we used the same system prompt in all our experiments:

“You are a highly intelligent and accurate medical domain named-entity recognition (NER) system.”

We experimented with the above system prompt and 4 different variations of the instruction string. For each instruction string, we computed the performance on the CONLL 2003 dataset (**Supplementary Table 3**). In our earlier iterations of the instruction string we requested the model to return medical entities in addition to PII entities. We expected this to improve the precision (e.g. drug “names” would fall under the medical category and would not be misclassified as a person’s name). However, this improvement was beneficial anecdotally in the earlier versions of GPT-3.5 and not for GPT-4. It was therefore discontinued.

**Supplementary Table 3.** Different variations of the instruction prompt attempted: We experimented with 4 different instruction prompts. “Prompt #4” is our best-performing prompt string on the CONLL 2003 dataset with an F1-Score of 0.87.

| Prompt Number | Instruction String | Precision | Recall | F1-Score |
|---|---|---|---|---|
| #1 | In the following text, extract all substrings containing the first names of people (key "FIRST_NAME"), last names of people (key "LAST_NAME"), middle names and initials of people (key "MIDDLE_NAME"), human ages (key "AGE"), IDs of medical records (key "MED_ID"), IDs of documents (key "DOC_ID"), calendar dates (key "DATE"), cities (key "CITY"), states (key "STATE"), street addresses (key "STREET") and countries (key "COUNTRY") as a JSON. Return substrings such as dates in every format in which they appear in the text. | 0.96 | 0.75 | 0.84 |
| #2 | In the following text, extract all substrings containing the first names of people (key "FIRST_NAME"), last names of people (key "LAST_NAME"), middle names and initials of people (key "MIDDLE_NAME"), email addresses (key "EMAIL"), human ages (key "AGE"), IDs of medical records (key "MED_ID"), IDs of documents (key "DOC_ID"), other alphanumeric IDs (key "ID"), calendar dates (key "DATE"), contracted or full names of days of the week (key "DAY"), contracted or full names of months (key "MONTH"), years (key "YEAR"), phone numbers (key "PHONE"), cities (key "CITY"), states (key "STATE"), street addresses (key "STREET"), countries (key "COUNTRY"), zip codes (key "ZIP"), names of hospitals (key "HOSPITAL"), drugs (key "DRUG"), diseases (key "DISEASE"), procedures (key "PROCEDURE"), and professions (key "PROFESSION") as a JSON. Return substrings in every format in which they appear in the text. | 0.91 | 0.79 | 0.85 |
| #3 | Following is a specification to list all medical terms and personal identifiers from provided text into JSON format where each key corresponds to an array of all terms from the text that are assigned the key. Medical terms like drugs(e.g sitagliptin), diseases and conditions(e.g NSCLC or non small cell lung cancer), procedures (e.g ACL knee reconstruction), lab tests(e.g Potassium levels) , medical devices, ICD, CPT, LOINC codes should be listed under a "MEDICAL" key. Names of people including doctors, nurses, and medical professionals should be listed under a "NAME" key. Dates, months, years, time and timestamp formatted text should be listed under a "DATE" key. Addresses, street names, zip codes, countries, counties and cities should be listed under a "Location" key. Organizations like hospital names, clubs, corporations, company names should all be listed under an "ORGANIZATION" key. Phone numbers , medical IDs, document IDs, database IDs, any numerical pattern that looks like an ID should be listed under an "ID" key. Email addresses should be listed under an "EMAIL" key. Similarly urls should be listed under a "URL" key. People's ages should be listed under an "AGE" key. Please use the preceding specification to apply on the following text: | 0.77 | 0.97 | 0.86 |
| #4 | In the following text, extract all substrings containing the following entity types as a JSON:<br>- First names of people (key "FIRST_NAME")<br>- Last names of people (key "LAST_NAME")<br>- Middle names and initials of people (key "MIDDLE_NAME")<br>- Human ages (key "AGE")<br>- IDs of medical records (key "MED_ID")<br>- IDs of documents (key "DOC_ID")<br>- Calendar dates (key "DATE")<br>- Cities (key "CITY")<br>- States (key "STATE")<br>- Street addresses (key "STREET")<br>- Countries (key "COUNTRY")<br>- Zip codes (key "ZIPCODE")<br>- Names of hospitals and other organizations (key "ORGANIZATION")<br>- Email addresses (key "EMAIL")<br>- Phone numbers (key "PHONE")<br>Return substrings such as dates in every format in which they appear in the text. | 0.81 | 0.95 | 0.87 |

#### Link to GitHub repo

https://github.com/lumenbiomics/udeid_llm

Deployment and Execution of Open-Source LLMs and GPT-3.5/GPT-4

1. Model Download Links:

- Meta-Llama-3-8b-Instruct: https://huggingface.co/meta-llama/Meta-Llama-3-8B-Instruct
- Meta-Llama-3-70b-Instruct: https://huggingface.co/meta-llama/Meta-Llama-3-70B-Instruct
- Mixtral-8x7B-Instruct-v0.1: https://huggingface.co/mistralai/Mixtral-8x7B-Instruct-v0.1
2. Infrastructure for Llama and Mixtral Models:

- Utilized Google Cloud Platform, specifically the G2 series:

○ Specifications: 96 cores, 384 GB RAM, and 8 x Nvidia L4 GPUs.
○ Approximately 1TB of disk space was allocated for storing the model files.
- Environment Setup:

○ Python version 3.8.10.
○ Deployed models using the <u>vLLM</u> library (Version 0.2.7).
○ Additional configuration:

■ Set tensor_parallel_size as 8 for all models.
■ For LLama3 70B, gpu_memory_utilization was set to 0.80 to avoid Out of Memory (OOM) errors.
3. Infrastructure for GPT-3.5 and GPT-4:

- Utilized Google Cloud Platform, E2 Series machine:

○ Specifications: 2 cores and 4GB RAM.
- Environment Setup:

○ Python version 3.8.10.
○ Deployed using the OpenAI Python library (Version 1.3.8) for GPT-3.5 and GPT-4 API calls.
- Specifically, the “gpt-3.5-turbo” and “gpt-4” model labels were used (https://platform.openai.com/docs/models/). The evaluation on the CONLL-2003 dataset was performed on 7th May 2024.

#### Number of failed documents for each LLM

Our evaluation requires a JSON output. It is possible that the generated output may not comply with the JSON schema for a given input document. We consider these documents to have failed and do not include them in our evaluation. The number of failed documents for the different models and datasets is summarized below.

**Supplementary Table 4.** Number of failed documents for each methodology: Our method is not a generative model and therefore does not produce unformatted output. We also find that GPT-4 produces consistently formatted JSON for all documents in the CONLL 2003 test set

| **Method** | **# failed documents (I2B2 2014 Test Set; N=514)** | **# failed documents (CONLL 2003 Test Set; N=217)** |
|---|---|---|
| Llama-3-8B | 409 | 26 |
| Llama-3-70B | 0 | 1 |
| Mixtral 8 x 7B | 191 | 56 |
| GPT-3.5 Zero Shot | N/A | 11 |
| GPT-3.5 One Shot | N/A | 2 |
| GPT-4 Zero Shot | N/A | 0 |
| GPT-4 One Shot | N/A | 0 |
| Locally augmented ensemble | **0** | **0** |

#### Examples of ChatGPT-4 errors

In this section, we provide an example input submitted to ChatGPT along with the prompt string to showcase false negatives errors (i.e. terms that are labeled as PII but not detected by ChatGPT). This example highlights how ChatGPT may underperform in structured evaluations with real-world examples while performing well anecdotally.

The following query was made using the OpenAI ChatGPT web application (https://chat.openai.com/) on March 6th, 2024.

Complete one-shot input string to ChatGPT-4:

~~~
In the following text, extract all substrings containing the following entity types as a JSON:
- First names of people (key “FIRST_NAME”)
- Last names of people (key “LAST_NAME”)
- Middle names and initials of people (key “MIDDLE_NAME”)
- Human ages (key “AGE”)
- IDs of medical records (key “MED_ID”)
- IDs of documents (key “DOC_ID”)
- Calendar dates (key “DATE”)
- Cities (key “CITY”)
- States (key “STATE”)
- Street addresses (key “STREET”)
- Countries (key “COUNTRY”)
- Zip codes (key “ZIPCODE”)
- Names of hospitals and other organizations (key “ORGANIZATION”)
- Email addresses (key “EMAIL”)
- Phone numbers (key “PHONE”)
Return substrings such as dates in every format in which they appear in the text.
Input text:
EU rejects German call to boycott British lamb. Peter Blackburn BRUSSELS 1996-08-22 The European Commission said on Thursday it disagreed with German advice to consumers to shun British lamb until scientists determine whether mad cow disease can be transmitted to sheep. Germany’s representative to the European Union’s veterinary committee Werner Zwingmann said on Wednesday consumers should buy sheepmeat from countries other than Britain until the scientific advice was clearer. “ We do n’t support any such recommendation because we do n’t see any grounds for it, “ the Commission’s chief spokesman Nikolaus van der Pas told a news briefing. He said further scientific study was required and if it was found that action was needed it should be taken by the European Union. He said a proposal last month by EU Farm Commissioner Franz Fischler to ban sheep brains, spleens and spinal cords from the human and animal food chains was a highly specific and precautionary move to protect human health. Fischler proposed EU-wide measures after reports from Britain and France that under laboratory conditions sheep could contract Bovine Spongiform Encephalopathy-LPR-BSE
-RPR- -- mad cow disease. But Fischler agreed to review his proposal after the EU’s standing veterinary committee, mational animal health officials, questioned if such action was justified as there was only a slight risk to human health. Spanish Farm Minister Loyola de Palacio had earlier accused Fischler at an EU farm ministers’ meeting of causing unjustified alarm through “ dangerous generalisation. “ “
Output JSON Response:
{“ORGANIZATION”: [“EU”, “European”, “Commission”, “Union”], “FIRST_NAME”:
[“Peter”, “Werner”, “Nikolaus”, “Franz”, “Fischler”, “Loyola”], “LAST_NAME”:
[“Blackburn”, “Zwingmann”, “van”, “der”, “Pas”, “Fischler”, “de”, “Palacio”],
“CITY”: [“BRUSSELS”], “COUNTRY”: [“Britain”, “France”, “Germany”], “DATE”: [“1996-08-22”]}
Input text:
JAKARTA 1996-12-06 Canada’s Bre-X Minerals Ltd and Barrick Gold Corp are to continue negotiations to hammer out a partnership agreement to develop the spectacular Busang gold find in Indonesia, sources close to the talks said on Friday. “ The negotiations will be held both in Toronto and in Jakarta, “ one source, speaking on condition of anonymity, told Reuters. Another source said most of the key negotiators from both Bre-X and Barrick had returned to Toronto, but declined to say if there had been any progress in their negotiations. Both sources said Bre-X and Barrick did not hold talks on Thursday with Mines and Energy Ministry Secretary-General Umar Said, who is coordinating the negotiations over the Busang find in East Kalimantan. The first source also said Bre-X had until December 21 to submit to the Indonesian Mines and Energy Ministry a feasibility study on the central region of the Busang property, estimated to contain 2.6 million ounces of gold. The richest parts of the property to the north and south of the central region have been estimated by Bre-X to contain 57 million ounces of gold. “ Bre-X is expected to complete the feasibility report by December 16 and submit it to the government before the December 21 deadline, “ the source said. He said Bre-X would then formally seek the permission of the Indonesian government to begin construction to develop Busang’s central region, which might take up to two years. The source declined to say if there had been any progress in the talks between Bre-X and Barrick. “ This is a huge project … we are not selling furniture, and Bre-X has 13,000 shareholders to answer to, “ the source said. “ While there has been some agreement in principle on some issues, there are still others such as procedures and mechanisms that needed to be sorted out, “ he added. The source said no new deadline had been set by the Mines and Energy Ministry for Bre-X and Barrick to strike a deal. The Ministry had given the companies until December 4 to complete a partnership deal, and advised Bre-X to take a 25 percent stake and Barrick 75 percent to develop the property. “ As far as I am aware, there’s been no new deadline, “ the source said. The Ministry’s Umar said on Thursday that both Bre-X and Barrick had responded positively to a government letter recommending a 25-75 split in the Busang gold property. The government also wants 10 percent of the property. Umar said the government had yet to receive a formal reply from the companies. He had said earlier that if the two companies failed to reach a partnership agreement, the government would explore other ways to expedite development of the Busang find. Bre-X has a partnership deal with PT Panutan Duta of the Panutan Group run by President Suharto’s eldest son, Sigit Harjojudanto, under which Panutan would receive $ 40 million over 40 months plus a 10 percent stake Busang’s richest parts. Barrick has teamed up with a construction company in the Citra Group of Suharto’s eldest daughter, Siti Hardianti Rukmana, in what Barrick had said was a partnership “ to prepare us for a potential mining development project “. Honda RV exceeds sales target.
Output JSON Response:
~~~

Output response from ChatGPT4:

~~~
{
“ORGANIZATION”: [“Bre-X Minerals Ltd”, “Barrick Gold Corp”, “Mines and Energy Ministry”, “PT Panutan Duta”, “Panutan Group”, “Citra Group”],
”FIRST_NAME”: [“Umar”, “Sigit”, “Siti”],
“LAST_NAME”: [“Said”, “Harjojudanto”, “Hardianti”, “Rukmana”], “CITY”: [“JAKARTA”, “Toronto”],
“COUNTRY”: [“Canada”, “Indonesia”],
“DATE”: [“1996-12-06”, “December 21”, “December 16”, “December 4”], “STATE”: [“East Kalimantan”]
}
~~~

In the above response, the following terms are not returned in the ChatGPT-4 response (False Negatives):

1. Names: “Suharto” (2 instances present in the document)
2. Organization: “Indonesian Mines”, “Reuters” (1 instance each in the document)
3. Location: “Busang” (7 instances present in the document)

Note: (1) CONLL 2003 has an additional entity type (MISC) which was not considered for our analysis.

(2) There were no false positives in this example.

### Cost Estimation

1. G2 Series Machine Costs (Google Cloud):

- Configuration with 8 L4 GPUs: $8/hr.
- Configuration with a single GPU: $0.71/hr.
2. E2 Series CPU Machine Costs (for GPT-3.5 and GPT-4 API Calls):

- Rate: $0.03/hr.
- We used the tiktoken Python library (https://github.com/openai/tiktoken) to calculate costs incurred for GPT-3.5 and GPT-4, based on OpenAI’s pricing (https://openai.com/pricing).

○ GPT-3.5: $0.0005/1K tokens (Input) & $0.0015/1K tokens (Output)
○ GPT-4: $0.03/1K tokens (Input) & $0.06/1K tokens (Output)
- Server costs for E2 during the usage of GPT-3.5 and GPT-4 APIs were minimal.
3. Exclusions:

- We did not include data storage and data transfer costs in our estimate.

## Footnotes

1 “SNOMED” is a registered trademark of the International Health Terminology Standards Development Organisation (IHTSDO).

